# Prehospital scale to differentiate intracerebral hemorrhage from large-vessel occlusion patients: A prospective cohort study

**DOI:** 10.1101/2023.09.20.23295876

**Authors:** A Freixa-Cruz, G Jimenez-Jimenez, G Mauri-Capdevila, Y Gallego-Sánchez, A García-Díaz, R Mitjana Penella, M Paul-Arias, C Pereira-Priego, E Ruiz-Fernández, S Salvany-Montserrat, A Sancho-Saldaña, E San-Pedro-Murillo, E Saureu, D Vázquez-Justes, F Purroy

**Author notes:** **Corresponding author:** Francisco Purroy, MD, PhD. Stroke Unit. Department of Neurology. Professor of the Universitat de Lleida. IRBLleida. Hospital Universitari Arnau de Vilanova de Lleida. Avda Rovira Roure, 80. Lleida 25198, Spain. /Fax 973248754. The authors declare no conflicts of interest.

## Abstract

**Background and rationale:** Evaluating scales to detect large vessel occlusion (LVO) could aid in considering early referrals to a thrombectomy-capable center in the prehospital stroke code setting. Nevertheless, they entail a significant number of false positives, corresponding to intracranial hemorrhages (ICH), which could result in a delay in medical attention and potential harm. Our study aims to identify easily collectible variables for the development of a scale to differentiate patients with ICH from LVO in a prehospital context.

**Methods:** We conducted a prospective cohort study of stroke code patients between May 2021 and January 2023. Patients were evaluated with CT/CT-Angiography at arrival. We compared clinical variables and vascular risk factors between ICH and LVO patients to design a prehospital ICH screening scale (PreICH).

**Results:** Out of 989 stroke code patients, we included 190 (66.7%) LVO cases and 95 (33.3) ICH cases. In the multivariate analysis, headache (odds ratio [OR] 3.56; 1.50-8.43), GCS<8 (OR 8.19; 3.17-21.13), SBP>160mmHg (OR 6.43; 3.37-12.26) and male sex (OR 2.07; 1.13-3.80) were associated with ICH, while previous hypercholesterolemia (HCL) (OR 0.35; 0.19-0.65) with LVO. The scale design was conducted, assigning a score to each significant variable based on its specific weight: +2 points for SBP > 160, +1 points for headache, +1 points for male sex, +2 points for GCS<8, and -1 points for HCL. The area under the curve (AUC) was 0.82 (0.77-0.87). A score ≥4 exhibited a sensitivity of 0.10, a specificity of 0.99, a positive predictive value of 0.21, and a negative predictive value of 0.98.

**Conclusion:** We present the development of a prehospital scale to discriminate between ICH and LVO patients, utilizing easily detectable variables in the prehospital setting.

## INTRODUCTION

### Background and rationale

Stroke remains a significant cause of morbimortality nowadays.^1–3^ Depicted by the time is brain concept, a rapid assessment, and early optimal treatment are crucial, as the probability of a complete neurological recovery decreases with every delayed minute.^4–6^ In ischemic strokes due to large vessel occlusion (LVO), mechanical thrombectomy (MT) has shown better functional results than the best medical treatment.^7–9^ Nevertheless, endovascular treatment (EVT) is only available in some institutions that can be located at a significant distance from the patient, leading to doubts about which healthcare facility should be the best primary transfer: (I) referral to a thrombectomy-capable center (mothership model) or (II) initial transport to the nearest hospital (drip-and-ship model).^10^ Prehospital scales such as RACE, G-FAST or LAMS can assess the risk of LVO based on clinical criteria with adequate accuracy thresholds.^11–17^ Recently, the RACECAT randomized clinical trial found no significant differences between the two transport strategies in Catalonia nonurban code stroke patients.^18^ In addition, it also raises the importance of identifying patients with intracranial hemorrhage (ICH), as they have a worse outcome with the ‘drip and ship’ model.^19–23^

In this context, improving the prehospital scales’ accuracy seems necessary to prevent false positives and thus better distinguish patients with LVO from those with ICH.^24–30^ Following this argument, the recent AHA guidelines emphasize the importance of studying the impact of pathways designed for the detection of patients with LVO on ICH patients.^31^

We hypothesize that there are clinical variables that can be collected in prehospital care, which are associated with ICH and could enable the development of prehospital scales.

## METHODS

### Data availability statement

Requests for access to the data reported in this article will be considered by the corresponding author.

### Study design and setting

This observational cohort study was conducted in our center (Hospital Arnau de Vilanova de Lleida, Spain) between April 2021 and July 2023. The study was carried out in two phases: (I) finding prehospital predictors of ICH and (II) developing an ICH screening scale. Study data were collected prospectively during recruitment and managed using REDCap electronic data capture tools hosted at the Biomedical Research Institute of Lleida. We followed the STROBE (Strengthening the Reporting of Observational Studies in Epidemiology) statement for reporting this study.^32^

### Participants

The cohort included patients over 18 who attended our center’s emergency department after activating a stroke code and were finally diagnosed with either AIS due to LVO or ICH. LVO was defined as the presence of an occlusion in the proximal anterior circulation (intracranial internal carotid artery [ICA]: distal ICA or T occlusions, M1 or M2 occlusion or tandem occlusions) detected by computed tomographic angiography (CTA) performed upon their arrival to the emergency department. ICH was defined as observing an intracerebral hematoma on the computed tomographic (CT) scan. A senior radiologist, blinded to clinical features, established the presence of LVO or ICH.

The exclusion criteria were ischemic strokes without LVO visible in urgent neuroimaging, stroke mimics as final diagnosis and traumatic or non-intracerebral intracranial hemorrhages. The subjects without written informed consent were also excluded.

### Variables, data collection and measurement

In some cases, the stroke code was activated according to the director’s Plan for Cerebrovascular Disease of the Government of Catalonia (Spain) in the prehospital environment, and patients were transported to our hospital. In other cases, the emergency medical service activated the stroke code upon the patient’s arrival. Therefore, any patient with clinical suspicion of an acute stroke within 24 hours of symptom onset and previous good quality of life was susceptible to activation.

All the data was collected prospectively during the time of recruitment and was included in an anonymized datasheet. The baseline demographics (age, sex), previous vascular risk factors (VRF), and previous treatments (antihypertensives, antiplatelets, anticoagulation and statins) were collected. VRF included diabetes mellitus (DM), prior hypertension (HTN), hypercholesterolemia (HCL), chronic kidney disease (CKD), atrial fibrillation (AF), alcoholism, smoking, coronary heart disease (CHD), non-previous stroke and prior cognitive impairment. HTN was defined as a systolic blood pressure ≥140 mm Hg or diastolic blood pressure ≥90 mm Hg or current use of antihypertensive medications. Cigarette smoking was defined as present if the patient reported smoking cigarettes during the past 5 years. HCL was defined as a total cholesterol concentration ≥220 mg/dL or the current use of lipid-lowering agents. DM was defined by history of fasting glucose ≥126 mg/dL or the current use of hypoglycemic medication. History of diagnosed coronary artery disease, peripheral arterial disease, atrial fibrillation, and valvular heart disease were also recorded.

We recorded systolic and diastolic blood pressure (SBP/DBP) using standardized measuring equipment from the EMS upon the patient’s arrival. We also documented the characteristics of symptom onset, whether it was a witnessed or unwitnessed onset, the presence of headache, language, sensory or motor impairment, campimetry deficit, Glasgow Coma Scale (GCS) score, the occurrence of seizures, and the National Institutes of Health Stroke Scale (NIHSS) score. According to the GCS scale, patients were divided into two groups: those with a score of less than eight and those with a score of 8 or higher. Patients were treated with usual management care at all times. In the patients evaluated in the prehospital environment, the RACE scale was also documented.

### Sample size

The sample size was calculated based on the primary outcome and analyzed using the GRANMO (v. 7.12) calculator The sample size was calculated based on the primary outcome, including an alpha risk of 0.05, a bilateral contrast, a beta risk of 0.20, a 1:2 ratio between the ICH and LVO groups, and estimated event proportions of 0.20 and 0.025, respectively. The calculation was performed using Granmo calculator (v. 7.12) webside (https://www.imim.es/ofertadeserveis/software-public/granmo/). The required minimum calculated sample size was 252 subjects, distributed among 168 in the LVO group and 84 in the ICH group. However, additional patients were included to facilitate the analysis of secondary outcomes, which may require a higher number of observations.

### Statistical analysis

The statistical analysis was conducted at the end of the study between June and July 2023 by a researcher not involved in the care process or the data collection.

A descriptive analysis of our sample was performed, including baseline demographics, pathological history, and previous treatments. Results were reported using mean and standard deviation (SD) or median and interquartile range (IQR) values. Categorical variables were presented as frequencies. We compared the baseline characteristics, the distribution of VRF, and the stroke severity between LVO and ICH patients. Student’s t-test and Mann-Whitney U test were employed to compare continuous variables based on their normal distribution. The Chi-squared test was used for categorical variables, and Fisher’s exact test was used when the expected cell frequency was <5. For the comparison of paired data, the Student’s t-test for paired samples and the Wilcoxon test were used to compare quantitative variables, depending on their parametric or non-parametric nature, respectively. McNemar’s test was applied to compare paired qualitative variables. A logistic multivariable analysis was conducted to identify independent risk factors associated with ICH, including significant (p<0.05) univariate variables. A score was assigned to each significant variable based on its specific weight to construct a predictive scale for assessing the risk of ICH. Lastly, the scale’s accuracy was evaluated by analyzing the area under the receiver operating characteristic curve (AUC). Subjects with missing data were not included in the statistical analysis. SPSS statistical package (version 25.0, SPSS, Chicago, IL, USA) was used to perform the statistical analysis. All tests were two-sided and conducted at a significance level of 0.05.

### Standard protocol approvals, registrations, and patient consents

All the subjects or legal representatives received the information related to the study and informed consent was obtained from all the participants. The study was approved by our local ethics committee “Comitè d’Ètica i Investigació Clínica de l’Hospital Universitari Arnau de Vilanova de Lleida” as a subproject of the program OMIC is BRAIN (acceptance code 2343).

## RESULTS

### Baseline characteristics

Out of 989 code stroke patients activated and evaluated in our hospital, 285 (28.8%) subjects were finally enrolled (45.1% female; mean age 75.1, SD 14.0). Among them, 190 (66.6%) were identified as LVO, while 95 (33.3%) were ICH (figure 1). In 171 (60%) patients, there was ambulance transportation with prior notice to the hospital.

**Figure 1.**
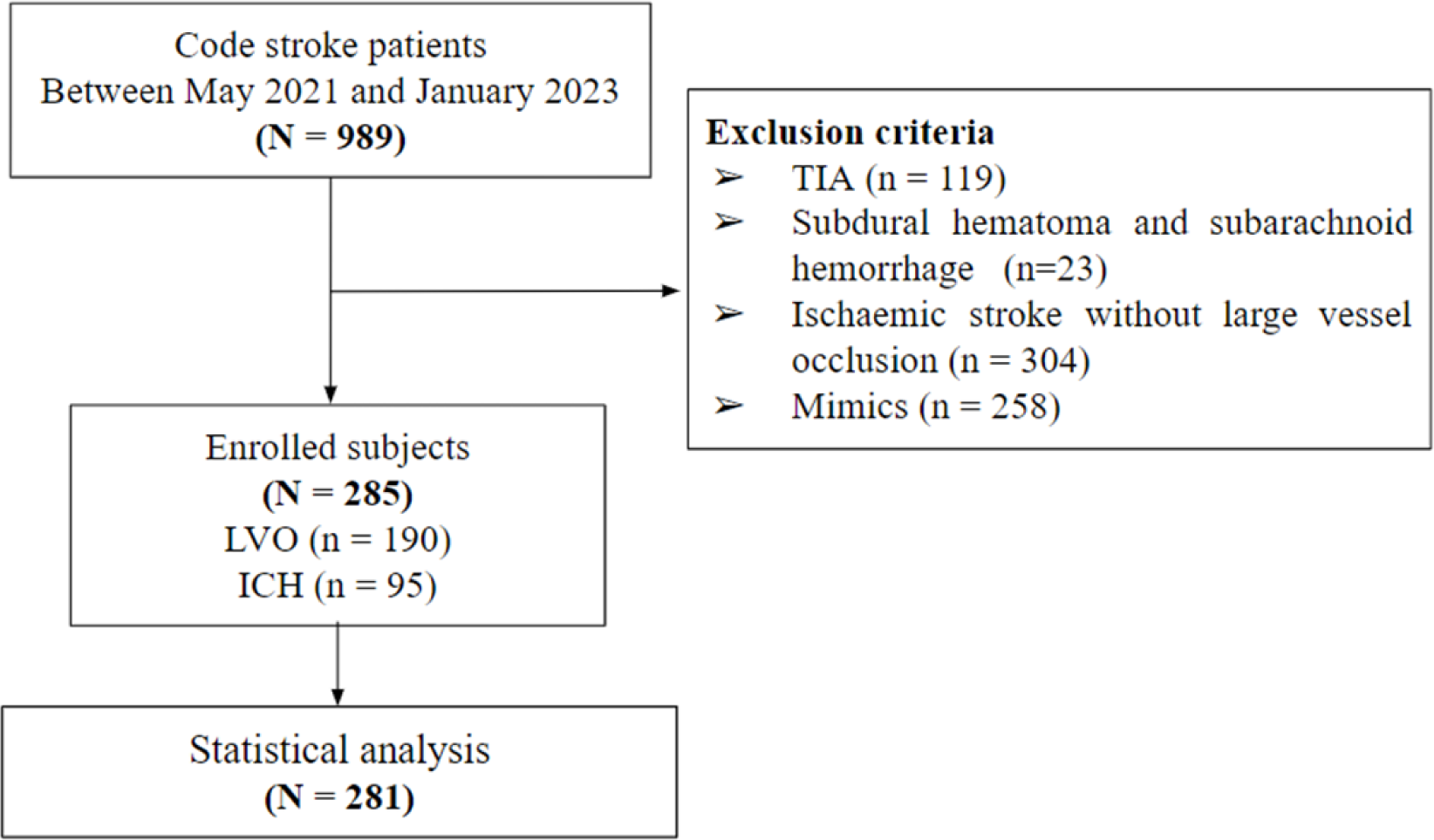
Flow-Chart.

### Differential variables among groups of patients

We observed a significant higher proportion of women, DM, DLD and AF in the LVO group (table 1). No significant differences were observed in the proportion of previous treatments and the event’s severity, as measured by the NIHSS, between the two groups. In contrast, SBP, DBP, the proportion of headache, GCS<8, and the onset of epileptic seizures were higher among ICH patients. The prevalence of visual field deficits was higher among the group with LVO. The RACE scores exhibited no significant difference between groups.

**Table 1.**
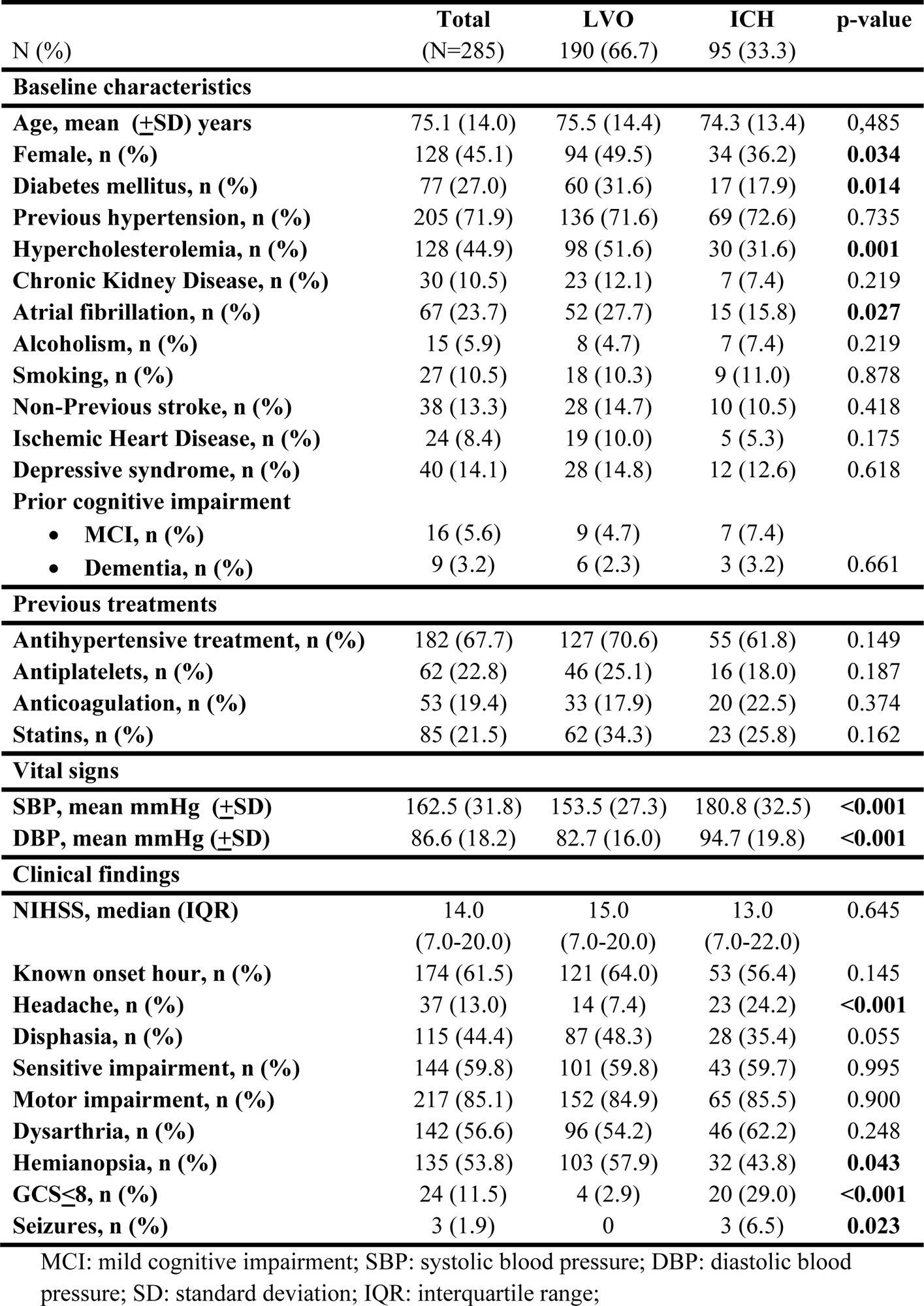
Baseline characteristics, previous treatments, vital signs and clinical findings.

|  | Total<br>(N=285) | LVO<br>190 (66.7) | ICH<br>95 (33.3) | p-value |
| --- | --- | --- | --- | --- |
| **Baseline characteristics** |  |  |  |  |
| Age, mean (±SD) years | 75.1 (14.0) | 75.5 (14.4) | 74.3 (13.4) | 0.485 |
| Female, n (%) | 128 (45.1) | 94 (49.5) | 34 (36.2) | **0.034** |
| Diabetes mellitus, n (%) | 77 (27.0) | 60 (31.6) | 17 (17.9) | **0.014** |
| Previous hypertension, n (%) | 205 (71.9) | 136 (71.6) | 69 (72.6) | 0.735 |
| Hypercholesterolemia, n (%) | 128 (44.9) | 98 (51.6) | 30 (31.6) | **0.001** |
| Chronic Kidney Disease, n (%) | 30 (10.5) | 23 (12.1) | 7 (7.4) | 0.219 |
| Atrial fibrillation, n (%) | 67 (23.7) | 52 (27.7) | 15 (15.8) | **0.027** |
| Alcoholism, n (%) | 15 (5.9) | 8 (4.7) | 7 (7.4) | 0.219 |
| Smoking, n (%) | 27 (10.5) | 18 (10.3) | 9 (11.0) | 0.878 |
| Non-Previous stroke, n (%) | 38 (13.3) | 28 (14.7) | 10 (10.5) | 0.418 |
| Ischemic Heart Disease, n (%) | 24 (8.4) | 19 (10.0) | 5 (5.3) | 0.175 |
| Depressive syndrome, n (%) | 40 (14.1) | 28 (14.8) | 12 (12.6) | 0.618 |
| **Prior cognitive impairment** |  |  |  |  |
| • MCI, n (%) | 16 (5.6) | 9 (4.7) | 7 (7.4) |  |
| • Dementia, n (%) | 9 (3.2) | 6 (2.3) | 3 (3.2) | 0.661 |
| **Previous treatments** |  |  |  |  |
| Antihypertensive treatment, n (%) | 182 (67.7) | 127 (70.6) | 55 (61.8) | 0.149 |
| Antiplatelets, n (%) | 62 (22.8) | 46 (25.1) | 16 (18.0) | 0.187 |
| Anticoagulation, n (%) | 53 (19.4) | 33 (17.9) | 20 (22.5) | 0.374 |
| Statins, n (%) | 85 (21.5) | 62 (34.3) | 23 (25.8) | 0.162 |
| **Vital signs** |  |  |  |  |
| SBP, mean mmHg (±SD) | 162.5 (31.8) | 153.5 (27.3) | 180.8 (32.5) | **<0.001** |
| DBP, mean mmHg (±SD) | 86.6 (18.2) | 82.7 (16.0) | 94.7 (19.8) | **<0.001** |
| **Clinical findings** |  |  |  |  |
| NIHSS, median (IQR) | 14.0<br>(7.0-20.0) | 15.0<br>(7.0-20.0) | 13.0<br>(7.0-22.0) | 0.645 |
| Known onset hour, n (%) | 174 (61.5) | 121 (64.0) | 53 (56.4) | 0.145 |
| Headache, n (%) | 37 (13.0) | 14 (7.4) | 23 (24.2) | **<0.001** |
| Disphasia, n (%) | 115 (44.4) | 87 (48.3) | 28 (35.4) | 0.055 |
| Sensitive impairment, n (%) | 144 (59.8) | 101 (59.8) | 43 (59.7) | 0.995 |
| Motor impairment, n (%) | 217 (85.1) | 152 (84.9) | 65 (85.5) | 0.900 |
| Dysarthria, n (%) | 142 (56.6) | 96 (54.2) | 46 (62.2) | 0.248 |
| Hemianopsia, n (%) | 135 (53.8) | 103 (57.9) | 32 (43.8) | **0.043** |
| GCS≤8, n (%) | 24 (11.5) | 4 (2.9) | 20 (29.0) | **<0.001** |
| Seizures, n (%) | 3 (1.9) | 0 | 3 (6.5) | **0.023** |

MCI: mild cognitive impairment; SBP: systolic blood pressure; DBP: diastolic blood pressure; SD: standard deviation; IQR: interquartile range;

### Multivariate analysis and scale design

Figure 2 shows the multivariate logistic regression analysis. Headache (odds ratio [OR] 3.56, 1.50-8.43, p=0.004), GCS<8 (OR 8.19, CI 3.17-21.13, p<0.001), SBP>160mmHg (OR 6.43, CI 3.37-12.26, p<0.001) and male sex (OR 2.07, 1.13-3.80, p=0.019) were associated with ICH, while HCL (OR 0.35, 0.19-0.65, p=<0.001) with LVO.

**Figure 2.**
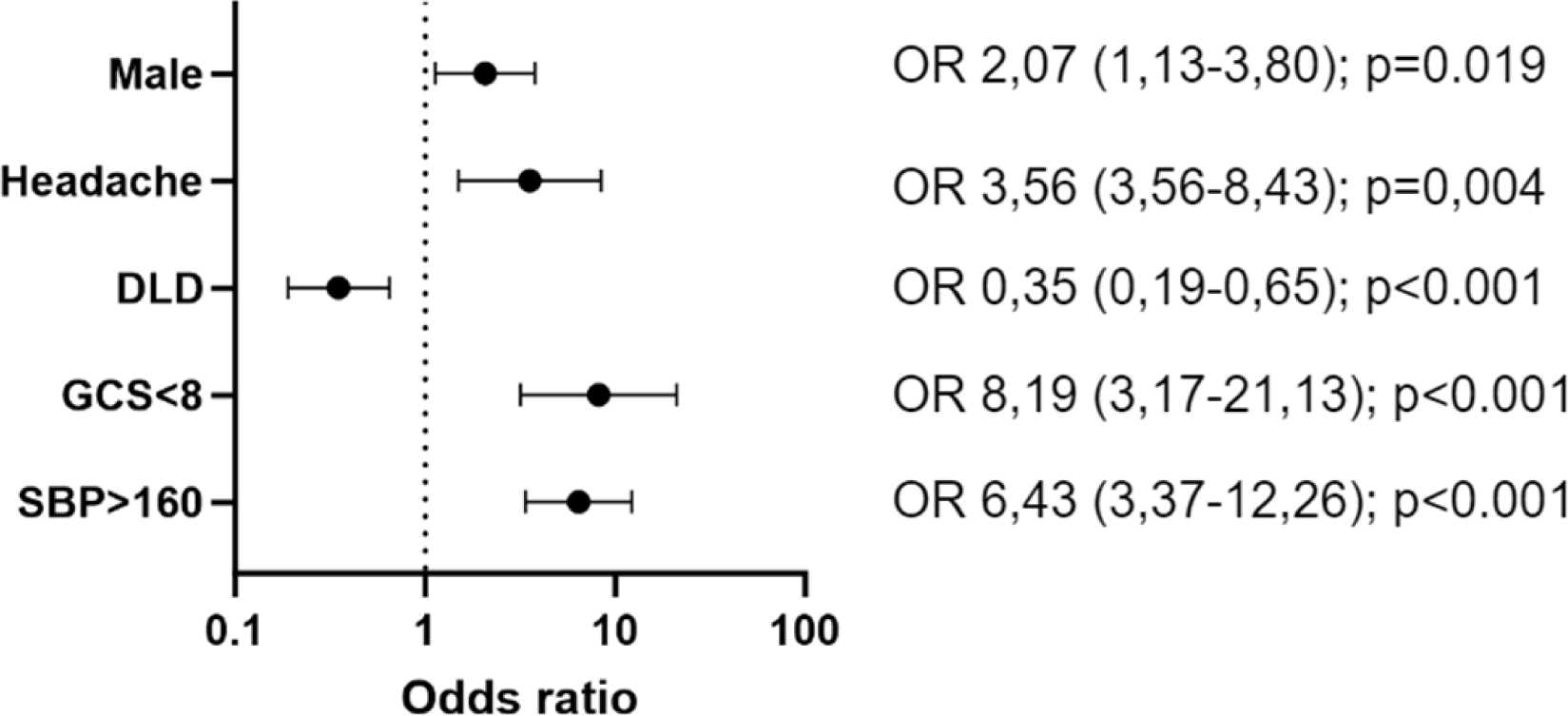
Multivariable analysis.

Based on these results, we designed a clinical scale which we named PreICH (prehospital clinical scale related to ICH). PreICH assigned a score to each significant variable based on its specific weight: +2 points for SBP > 160, +1 points for headache, +1 points for male gender, +2 points for GCS<8, and -1 points for HCL. The scale could be calculated in 281 (98.6%) patients. Figure 3 shows the distribution of patients according to the score spectrum, observing the highest proportion in 2 points (24.9%) followed by 1 and 0 points (23.1% and 19.2%, respectively). Figure 4 depicts the distribution of patients with ICH and LVO based on the potential scale values. At the lowest scores (-1, 0, and +1), there was a notable proportion of LVO cases compared to ICH cases. Conversely, this trend was reversed at the highest scores (+4, +5, +6). Notably, all patients with a score of -1 had LVO, while all those with a score of +6 presented ICH.

**Figure 3.**
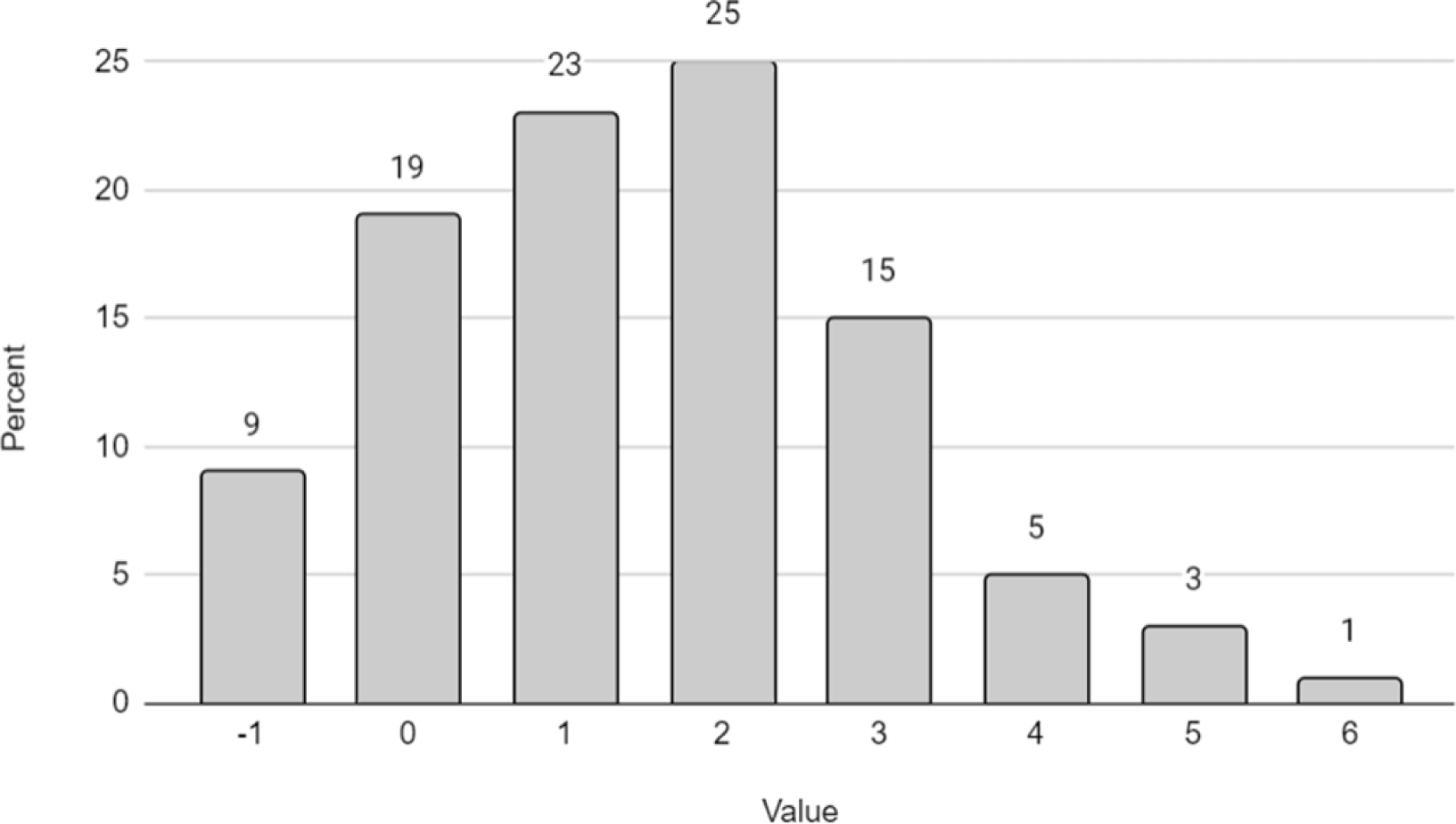
Distribution of subjects by PreICH scores.

**Figure 4.**
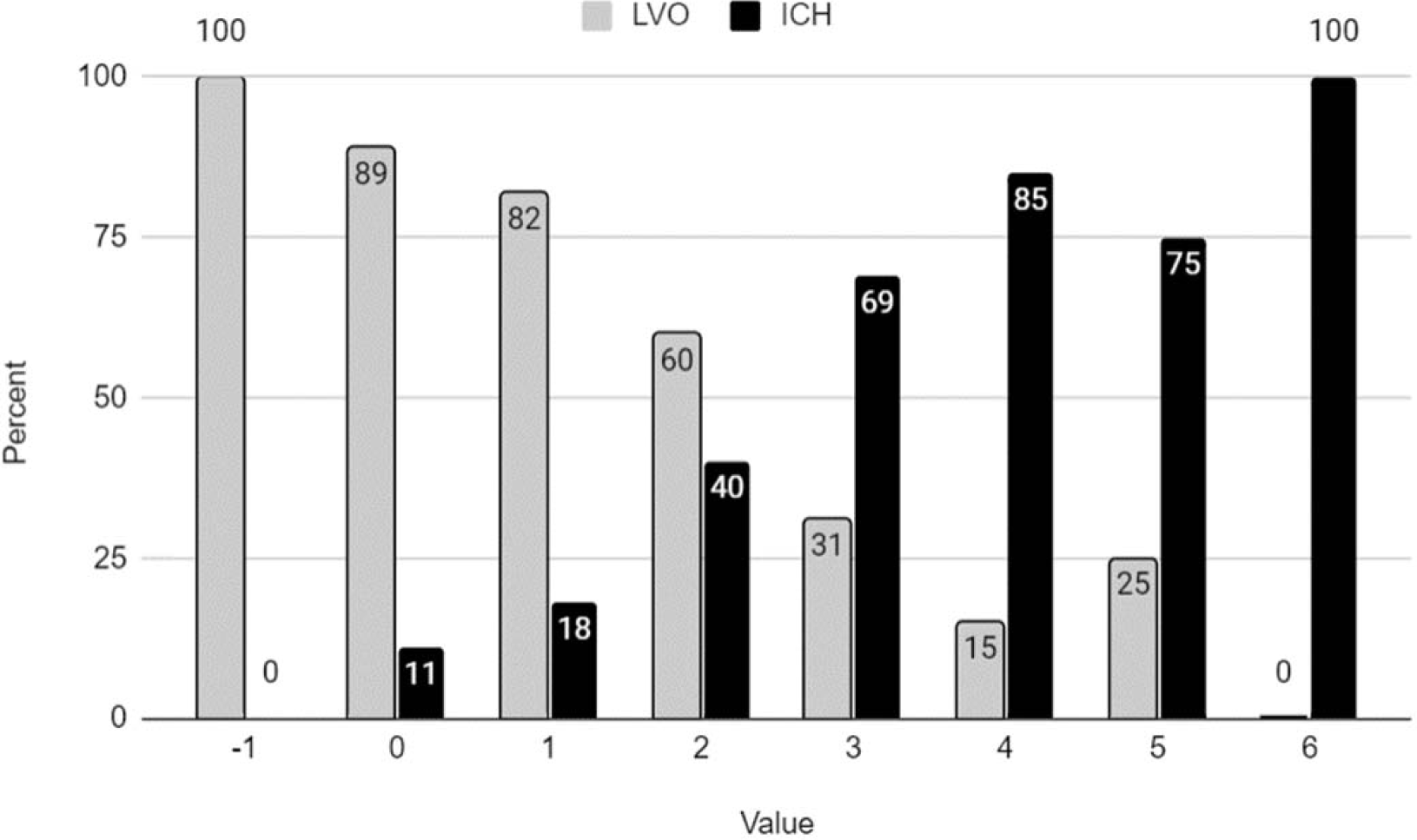
Distribution of ICH and LVO subjects by PreICH scores.

The area under the curve (AUC) analysis (Figure 5) was 0.82 (0.77-0.87) p<0.001, with the optimal cutoff at 4 points. A PreICH score ≥4 exhibited a sensitivity of 0.10, a specificity of 0.99, a positive predictive value of 0.21, and a negative predictive value of 0.98 (table 2).

**Figure 5.**
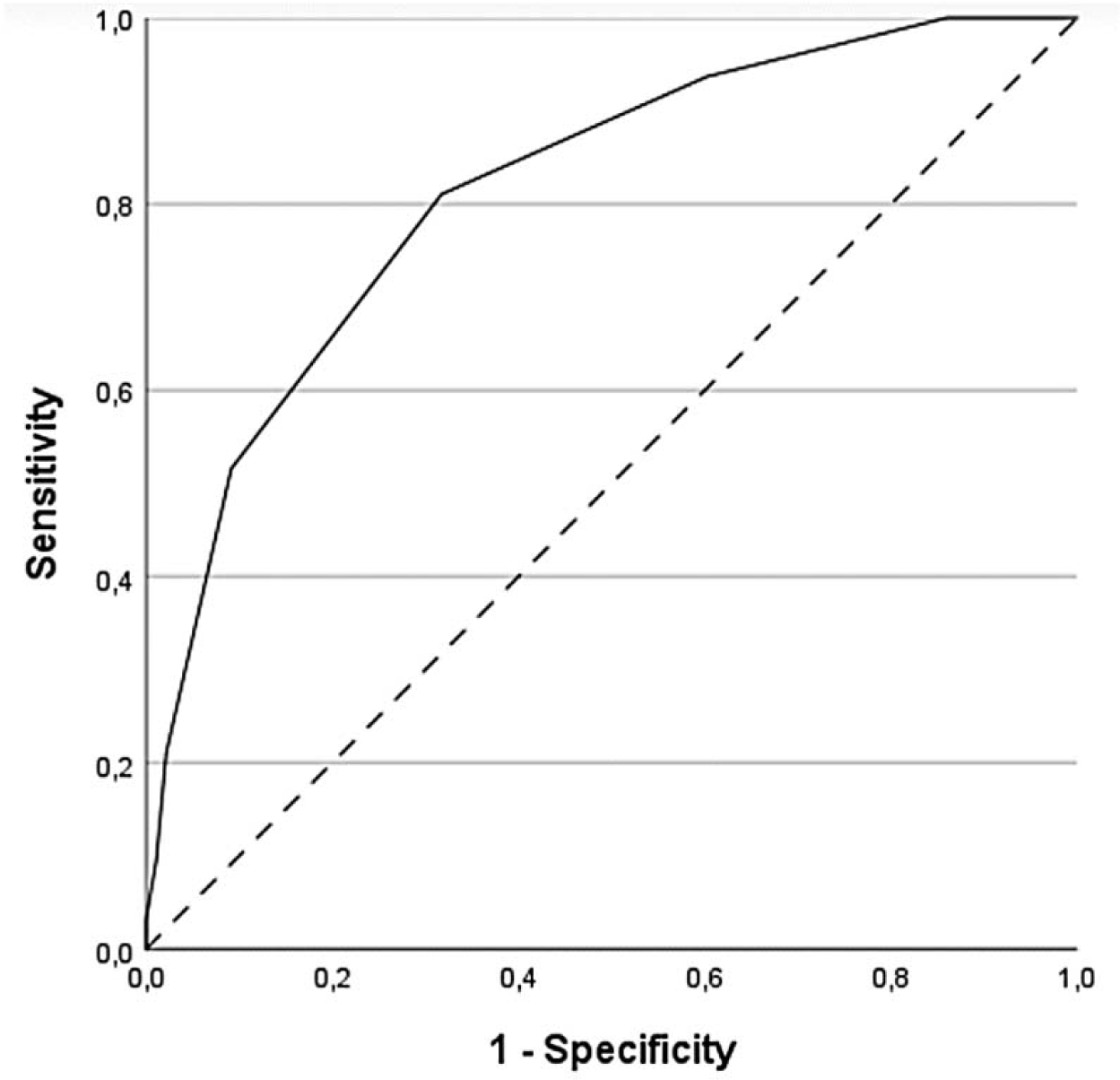
PreICH area under the curve (AUC) analysis.

**Table 2.** Distribution of ICH and LVO subjects by PreICH >4 cut-off.

|  | **LVO**<br>**N=186 (66.2)** | **ICH**<br>**N=95 (33.8)** | **Total** |
| --- | --- | --- | --- |
| **PreICH <4, N (%)** | 182 (97.8) | 75 (78.9) | 257 (91.5) |
| **PreICH $\geq 4$, N (%)** | 4 (2.2) | 20 (21.1) | 24 (8.5) |
|  | 186 (66.2) | 95 (33.8) |  |

Table 3 displays the distribution of patients with ICH and LVO based on significant scoring on the RACE and PreICH scales. In our cohort, 100 patients had RACE scores of ≥5. Of those, 35 had ICH. Among those with RACE score of ≥5, only 8 had PreICH scores of ≥4. Out of these 8 patients, 7 (87.5%) of them had ICH. Globally, 13 (7.6%) patients had PreICH score of ≥4. In 11 (84.6%) of them, ICH was confirmed.

**Table 3.** Distribution of ICH and LVO subjects depending on significant / non-significant scoring of the RACE and PreICH scales.

|  |  | **LVO**<br>N= 112 (65.5) | **ICH**<br>N=59 (34.5) | **Total**<br>N=171 |
| --- | --- | --- | --- | --- |
| **RACE<5**<br>N=71 (41.5) | **PreICH <4** | 47 (71.2) | 19 (28.8)) | 66 (38.6) |
|  | **PreICH ≥4** | 1 (20.0) | 4 (80.0) | 5 (2.9) |
| **RACE≥5**<br>N=100 (58.5) | **PreICH <4** | 63 (68.5) | 29 (31.5) | 92 (53.8) |
|  | **PreICH ≥4** | 1 (12.5) | 7 (87.5) | 8 (4.7) |

## DISCUSSION

### Key results and interpretation

We propose a straightforward preclinical scale to differentiate between ICH and LVO patients. This scale is based on variables that can be easily obtained during the initial evaluation of stroke code patients. Specifically, SBP, headache during the event, and a decreased level of consciousness were associated with ICH, while a history of previous HCL was linked to LVO. Additionally, we observed differences in terms of gender, with ICH being overrepresented among men.

The interest in prehospital scales in the field of stroke has gone hand in hand with the development of interventional therapies for LVO patients.^4,6–9^ The fact that the care of patients with LVO is centralized in tertiary stroke centers has raised the importance of identifying potential candidates for these therapies who could benefit from direct transportation to these centers.^10,19,20^ However, ICH patients experience worse outcomes if their hospital care is delayed, primarily due to them being more dynamic individuals who frequently undergo neurological deterioration.^21^

In this context, we believe that our scale provides value as long as we propose to be used in conjunction with scales aimed at identifying patients with LVO. Our scale emphasizes its specificity to ensure an adequate negative predictive value. This approach ensures that the identified patients have a higher probability of suffering from ICH and thus prevents LVO patients from being adversely affected by delays in accessing thrombectomy centers.

For years, the greater severity of ICH compared to ischemic stroke patients has been recognized. Thus, a higher likelihood of coma, vomiting, seizures, and blood pressure has been described. Therefore, our results would be consistent with the previous literature.^24–30^

To date, clinical scales aimed at enhancing the diagnosis of ICH focused on distinguishing this condition from other types of cerebrovascular disease like globally ischemic strokes, transient ischemic attack patients or stroke mimics rather than differentiating it from patients with LVO.^27–29^ Specifically, Ye et al., for example, distinguishes between ICH and LVO based on age and blood pressure. The ph-ICH score was based on SBP and level of consciousness, similar to our approach. However, the authors also considered the severity of neurological deficits.^30^ Interestingly, the Japan Urgent Stroke Triage (JUST) score is a clinical prediction rule to classify suspected stroke patients into different types, including LVO at the prehospital stage. Like in our cohort, the proportion of headache, male and high SBP was higher in ICH than in LVO patients. On the contrary, the authors did not take into account the history of HCL.^29^

### Limitations and generalisability

Our study has several limitations that are worth explaining. On the one hand, although a preliminary sample calculation was conducted and has been achieved and surpassed during the inclusion period, our work proposes a scale for ICH that needs validation. It is important to note that not all patients were able to report experiencing headache due to some having severe deficits or speech alterations. Similarly, it was not possible to determine whether seizures had occurred accurately in cases with no witness to the episode. On the other hand, we only recorded the presence of diplopia and not its intensity. Furthermore, in the case of a history of HCL, we considered the previous diagnosis primarily made by the general practitioner or the use of medications related to diabetes. There might be biases in the definition of HCL, especially considering its evolving definition over the years. In addition, another limitation is the choice of the cutoff point. In our study, we aimed to emphasize specificity, but validation in another cohort and application by paraclinical professionals would be necessary to determine its validity. Despite these limitations, we believe our results can be generalized, considering that the proportion of stroke code patients in the initial cohort who experienced ICH or LVO is similar to that of other cohorts.

## CONCLUSIONS

Our clinical scale is a simple proposal based on clinically assessable variables in the prehospital setting, which, when applied alongside other validated scales for detecting suspected LVO, can enhance the management pathways for patients with a final diagnosis of ICH. However, validation is necessary to standardize its clinical usage.

## Declaration of conflicting interests

Authors declare no conflict of interest

## Funding

This study was supported by the Government of Catalonia-Agència de Gestió d’Ajuts Universitaris i de Recerca (2021SGR01479); Instituto de Salud Carlos III and co-funded by European Union (ERDF/ESF, “Investing in your future” and “A way to build Europe”) (PI20/01575) and the RICORS Research Network (RD21/0006)

## Author contribution

FP and GJ conceived the study. FP and AF designed experiments. AF, GM, YG, AGD, MP, ER, SS, CP, AS, ES, D, ES cohorts’ recruitment and clinical data acquisition. RM reviewed neuroimaging. FA and AF sample processing and data analysis. FP and AF participated on data interpretation and draft the manuscript. All authors critically revised and approved the final version of the manuscript. FP procured funding.

## Non-standard Abbreviations and Acronyms

AF: Atrial fibrillation
AIS: Acute ischemic stroke
AUC: Area under the curve
CHD: Coronary Heart Disease
CI: Confidence interval
CKD: Chronic kidney disease
CSC: Comprehensive Stroke Center
CTA: Computed tomography angiography
DBP: Diastolic blood pressure
DM: Diabetes mellitus
EVT: Endovascular treatment
GCS: Glasgow coma scale
HCL: Hypercholesterolemia
HTN: Hipertension
ICH: Intracerebral hemorrhage
IQR: Interquartile range
LVO: Large vessel occlusion
MT: Mechanical thrombectomy
NIHSS: National Institutes of Health Stroke Scale
OR: Odds ratio
PSC: Primary Stroke Center
SBP: Systolic blood pressure
SD: Standard deviation
TIA: Transient ischaemic attack
VRF: Vascular risk factor

## Notes

### Competing Interest Statement

The authors have declared no competing interest.

### Clinical Trial

NA

### Author Declarations

CEIC Hospital Universitari Arnau de Vilanova de LLeida

